# Hearing Loss and Tinnitus: Uncover the Mechanism of Tinnitus using Resting-State Functional Magnetic Resonance Imaging (rs-fMRI) Technologies

**DOI:** 10.1101/2022.06.26.22276920

**Authors:** Haoliang Du, Xu Feng, Xiaoyun Qian, Jian Zhang, Bing Liu, Zhichun Huang, Xia Gao

## Abstract

**Purpose:** This project aimed to investigate the differences in the intra-regional brain activity and inter-regional functional connectivity in subjects with tinnitus only and subjects with hearing loss and tinnitus, using resting-state functional magnetic resonance imaging (rs-fMRI) technologies, including the Amplitude of Low-Frequency Fluctuations (ALFF), regional homogeneity (ReHo), and Voxel-Wise Functional Connectivity (FC).

**Method:** We acquired rs-fMRI scans from 82 subjects (21 tinnitus subjects without hearing loss, 32 subjects with tinnitus and hearing loss, and 29 subjects as healthy control). Age, gender, and year of education were matched across all three groups. We consecutively performed ALFF, ReHo, and Voxel-Wise Functional Connectivity (FC) for all subjects.

**Result:** Compared with the control group (CN), subjects with tinnitus only (T group) and with tinnitus and hearing loss (T+H group) manifested significantly reduced ALFF and ReHo activity within the left and right dorsolateral superior frontal gyrus (SFG). Additional Voxel-Wise Functional Connectivity (FC) revealed decreased connectivity between the dorsolateral SFG (left and right) and right Superior Parietal Gyrus (SPG), right Middle Frontal Gyrus (MFG), and left medial Superior Frontal Gyrus (mSFG) within these two groups. No significant differences were observed between the T and T+H groups.

**Conclusion:** Upon analyzing our data, we suggested disruptions in brain regions responsible for attention and stimuli monitoring and orientations contribute to tinnitus generation. Thus, hearing loss might not be the primary cause of tinnitus.

## 1 Introduction

Tinnitus is a phantom auditory perception without corresponding acoustic or mechanical correlations in the cochlea. Tinnitus represents one of the most common yet distressing otologic pathologies, and it affects approximately 10 to 15% of the adult population (Zenner et al., 2016; Minami et al., 2018). Existing studies reported that tinnitus is commonly associated with hearing loss, middle and inner ear-related pathologies, noise exposure, ototoxic medications and chemicals, aging, sleep deprivation, anxiety, depression, head and neck injuries, and temporomandibular joint (TMJ) dysfunction. In addition, due to its bothersome nature, tinnitus patients may perceive sleep deprivation, depression, anxiety, poor attention, decreased speech recognition, and even a reduced quality of life (Baguley et al., 2013).

Existing literature indicates that tinnitus involves multiple systems responsible for emotion, attention, memory, and functional executions. Thus, current research on tinnitus shifted to uncover the neural mechanisms behind tinnitus (Hu et al., 2021; Zenke et al., 2021). Kapolowicz and Thompson reported that tinnitus might be closely related to an imbalance between the auditory neuronal excitation and inhibition network, leading to plasticity changes in the central auditory system (Kapolowicz & Thompson, 2020). In research by Knipper et al. (2021), the researchers proposed that hearing loss may contribute to a top-down mechanism that leads to tinnitus perception (Knipper et al., 2021). Khan and colleagues suggested that tinnitus might be a compensatory response to damage to the peripheral hearing system (Khan et al., 2021). Cai, Xie, and colleagues reported abnormal functional connectivity in the auditory and non-auditory cortices in patients with hearing loss and tinnitus (Cai et al., 2020). Zhou and colleagues suggested that patients with hearing loss and tinnitus demonstrated abnormal intra-regional neural activity and disrupted connectivity in the hub regions of some non-auditory networks, including the default mode network (DMN), optical network, dorsal and ventral attention network (DAN & VAN), and central executive network (CEN) (Zhou et al., 2019). Finally, Minami and colleagues reported that tinnitus patients with hearing loss showed a statistically significant reduction in auditory-related functional connectivity than the control group (Minami et al., 2018). Given these points, the underlying neural mechanism of tinnitus generation remains unclear since the cause of tinnitus can be multifactorial (Zhang, 2013; Feng et al., 2018). In addition, the above findings cannot effectively elucidate the 20% of tinnitus patients with clinically normal hearing or the 50% of those with hearing loss who do not perceive tinnitus (Khan et al., 2021).

Since neuronal pathologies are less likely to generate significant morphological variations, molecular imaging techniques are essential for revealing such minor changes inside the brain. Vast development and advancement of molecular probes that bind biochemical markers and instrumentations have provided new insight into the human brain organization, visualization of brain structures, and brain-behavior relationships. Functional magnetic resonance imaging (fMRI) can yield non-invasive examination, localization, and lateralization of critical brain functions because of its widespread availability, non-invasive nature, relatively low cost, and adequate spatial resolution (Mier & Mier, 2015). Among all fMRI techniques, the resting-state functional magnetic resonance imaging (rs-fMRI) has been favored over other fMRI techniques due to its lack of difficulty in the signal acquisition, least amount of input required from the patients, and ability to identify the functional areas in different patient populations (Lv et al., 2018).

Resting-state fMRI has been widely performed in both healthy subjects and patients with tinnitus. As an effective and powerful tool for characterizing functional and network connectivity, rs-fMRI provides valuable information for a more comprehensive understanding of tinnitus’ neural mechanism (Chen & Glover, 2015; Lv et al., 2018). Compared to tasked-based fMRI imaging, test subjects do not need to perform specific tasks during the resting-state fMRI. Research has revealed that the low-frequency oscillations of the resting-state fMRI signal relate to spontaneous neural activity. The Amplitude of Low-Frequency Fluctuations (ALFF) and regional homogeneity (ReHo) are the primary tools for assessing functional segregation (Lee et al., 2012; Lv et al., 2018). The ALFF can detect the regional intensity of spontaneous fluctuations in the blood-oxygen-level-dependent (BOLD) signal, which defines the brain’s spontaneous neural activity in specific regions and physiological states. ReHo is a voxel-based measurement of brain activity that evaluates the synchronization or similarity between the time series of a given voxel and its nearest neighbors (Soares et al., 2016). Since ReHo does not require a priori definition of regions of interest (ROIs), it can provide information about the regional activity of destinated brain areas. However, while ALFF and ReHo reflect different aspects of regional neural activity, they can not reveal the functional connectivity between two destinated brain regions. On the other hand, voxel-wise functional connectivity (FC) describes the relationship between the neuronal activation patterns of anatomically separated brain regions and reflects functional communication between regions (Lee et al., 2012; Soares et al., 2016; Lv et al., 2018).

Hearing loss is commonly considered the cause of tinnitus, yet many patients can perceive tinnitus without hearing loss. Therefore, we aim to discover the generating mechanism for tinnitus among patients without hearing loss by investigating the differences in the intra-regional brain activity and inter-regional functional connectivity in subjects with tinnitus only and subjects with hearing loss and tinnitus, using ALFF, ReHo, and Voxel-Wise Functional Connectivity technologies.

## 2 Method

### 2.1 Subjects’ Clinical Information

The Research Ethics Committee of the Affiliated Zhongda Hospital of Southeast University approved this study. All individuals provided written informed consent before they participated in the study. We recruited eighty-two subjects (all right-handed, with at least eight years of education), including twenty-one tinnitus subjects without hearing loss (T), thirty-two tinnitus subjects with hearing loss (T+H), and twenty-nine healthy subjects as the control group (CN) through virtual recruitment promotions and community screening sessions from September 2011 to September 2013. The patients were group-matched in terms of age, sex and education. Twenty-five subjects perceived bilateral tinnitus, and the rest, twenty-eight subjects, perceived unilateral tinnitus.

We collected pure-tone hearing thresholds (PTA for 250, 500, 1000, 2000, 4000, 6000, and 8000 Hz) from all recruited subjects. Subjects with 7-frequency PTA below 25 dB HL were considered clinically normal hearing. In addition, we performed comprehensive tympanometry, diagnostic distortion-product otoacoustic emissions (DPOAE), and diagnostic auditory brainstem response (ABR) for all subjects to rule out middle ear pathologies and auditory neuropathy (ANSD). Furthermore, we collected crucial clinical information from all subjects, including the duration of tinnitus and the presence of insomnia.

To assess the severity and distress associated with tinnitus, we distributed the Iowa version of the tinnitus handicap questionnaire (THQ) (Kuk et al., 1990) to both the T and T+H groups. In addition, a 5-level visual analog scale (VAS) was used to measure the level of insomnia of the recruited participants (4 being constant insomnia to 0 being almost no insomnia). We also distributed the Self-Rating Depression Scale (SDS) and the Self-Rating Anxiety Scale (SAS) questionnaires to all subjects for anxiety and depression screening (Zung, 1986; Zung, 1971). Finally, we summarized the subjects’ characteristics for each group in table 1.

**Table 1.**
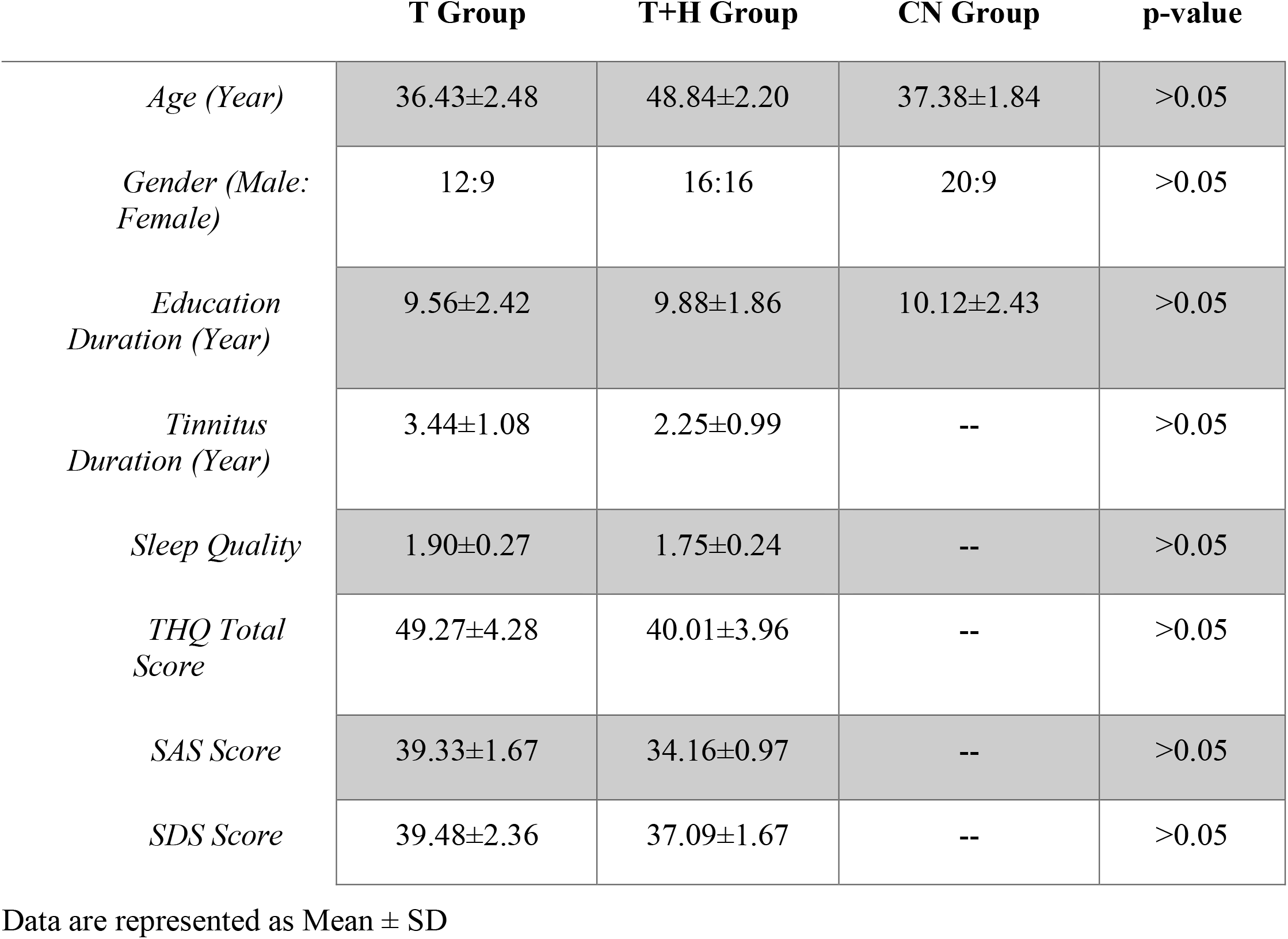
Subject Characteristics of the Tinnitus Group (T), Tinnitus with Hearing Loss Group (T+H), and Control Group (CN)

### 2.2 Subject Exclusion Criteria

The exclusion criteria for this study included Meniere’s disease, objective tinnitus, pulsatile tinnitus, histories of consuming alcohol, severe smoking, head and neck injuries, epilepsy, stroke, Alzheimer’s disease, Parkinson’s disease, cancer, MRI contraindications, primary psychiatric conditions including Generalized Anxiety Disorder (GAD), depression and Schizophrenia, and severe visual impairment. None of our subjects failed the depression and anxiety screening.

### 2.3 fMRI Scanning & Data Acquisition

We acquired the imaging data using a 3.0 T MRI scanner (Siemens MAGENETOM Trio, Erlangen, Germany) with a standard head coil. We provided all subjects with foam paddings and earmuffs to minimize head motion and noise exposure during the scanning process. The subjects were instructed to remain calm during the scan with their eyes closed without falling asleep or thinking of anything particular. Functional images were obtained axially using a gradient echo-planar sequence sensitive to BOLD contrast as follows: repetition time (TR) =2000 ms; echo time (TE) = 25 ms; slices = 36; thickness = 4 mm; gap = 0 mm; field of view (FOV) = 240 × 240 mm; acquisition matrix = 64 × 64; and flip angle (FA) = 90°.

### 2.4 Amplitude of Low-Frequency Fluctuations (ALFFs): Preprocessing & Analysis

Resting-state ALFF can reflect spontaneous neural activity and yield physiologically meaningful results. Pre-processing of the ALFF images was performed using the toolbox Data Processing Assistant for Resting-State fMRI (DPARSF 5.2), Statistical Parametric Mapping (SPM 12), and Matlab 2021b. We removed the first five volumes from each time series to account for subjects’ adaptation to the scanning environment. Slice timing and re-alignment for head-motion correction were performed for the remaining 175 images. Afterward, we performed the following procedures: spatially normalized into the stereotactic space of the Montreal Neurological Institute (MNI) (resampling voxel size = 3 × 3 ×3 mm^3^) and smoothed using a Gaussian kernel of 6 mm full width at half-maximum (FWHM), de-trending, and filtering (0.01–0.08 Hz). The subjects with a head motion with more than 2.0 mm displacement or a 2.0-degree rotation in the *x, y*, or *z* directions were excluded from this study.

We then analyzed the ALFF data by transforming time to the frequency domain using Fast Fourier Transform. Next, we computed the square root of the power spectrum and averaged squared across 0.01–0.08 Hz at each voxel. The calculated averaged square root was taken as the ALFF. Finally, the ALFF of each voxel was divided by the global mean ALFF value for standardization.

### 2.5 Regional Homogeneity (ReHo): Preprocessing & Analysis

The ReHo calculates the synchronization of low-frequency fluctuations between a given voxel and neighboring voxels, reflecting the neural function synchronization in the local brain region. Pre-processing of ReHo images was performed using the toolbox Data Processing Assistant for Resting-State fMRI (DPARSF 5.2), Statistical Parametric Mapping (SPM 12), and Matlab 2021b. We removed the first five volumes from each time series to account for subjects’ adaptation to the scanning environment. Slice timing and re-alignment for head-motion correction were performed for the remaining 175 images. The following procedures were performed: spatially normalized into the stereotactic space of the Montreal Neurological Institute (MNI) (resampling voxel size = 3 × 3 ×3 mm^3^), de-trending, and filtering (0.01–0.08 Hz).

After the pre-processing stage, we performed the image calculation using the Kendall coefficient of concordance of the time series of a given voxel with its 27 nearest neighbors. Next, ReHo analyses were calculated using the DPARSF 5.2 software. The ReHo value of each voxel was then standardized by partitioning the primal value using the global mean ReHo value. Finally, the data were smoothed with a Gaussian kernel of 6 mm full-width at half maximum (FWHM) for further statistical analysis.

### 2.6 Voxel-Wise Functional Connectivity (FC): Pre-processing & Analysis

We performed the Voxel-Wise FC analysis using the toolbox Data Processing Assistant for Resting-State fMRI (DPARSF 5.2), Statistical Parametric Mapping (SPM 12), and Matlab 2021b. The first ten volumes were removed from each time series to account for subjects’ time to adapt to the scanning environment. Then, slice timing and re-alignment for head-motion correction were performed for the remaining 170 images. Afterward, the procedures were carried out as follows: spatially normalized into the stereotactic space of the Montreal Neurological Institute (MNI) (resampling voxel size = 3 × 3 ×3 mm^3^) and smoothed using a Gaussian kernel of 6 mm full width at half-maximum (FWHM), de-trending, and filtering (0.01–0.08 Hz). Subjects with a head motion with more than 2.0 mm displacement or a 2.0-degree rotation in the *x, y*, or *z* directions were excluded.

We extracted the ALFF and ReHo differences in brain regions between tinnitus subjects without hearing loss and with hearing loss for Voxel-Wise FC analysis and defined them as seeds. We then used the average time series of seeds as a reference and calculated the Pearson correlation coefficient between the average signal change of each seed and the time sequences of other voxels in the brain. Finally, we converted the correlation coefficient to a z-value using Fisher’s z-transformation.

### 2.7 Statistical Analysis and Graphic Illustration

The one-way analysis of variance (ANOVA) was firstly conducted to test mean differences in ALFF, ReHo, and functional connectivity (FC) between the control group (CN), the group with tinnitus only (T), and the group with both hearing loss and tinnitus (T+H) (Matlab 2021b). The statistically-significant difference between the groups was determined at p < 0.05. Subjects’ age and gender were included as nuisance covariates. Next, we applied Family-Wise Error (FWE) correction for multiple comparisons, using voxel-level inference at p < 0.001 and cluster-level inference at p<0.05. Two-sample t-tests were then conducted to investigate the ALFF, ReHo, and functional connectivity (FC) differences between subjects with tinnitus only (T) and control group (CN), subjects with tinnitus and hearing loss (T+H) and control group (CN), and subjects with tinnitus only (T) and subjects with tinnitus and hearing loss (T+H). The statistically-significant difference between the groups was determined at p < 0.05. Finally, we used the MRIcroGL software to draw 3-dimensional brain images to display the brain areas with statistically significant differences.

## 3. Results

### 3.1 ALFF Results

We discovered significant ALFF value differences in the left and right dorsolateral SFG for both T and T+H groups compared to the CN group (Fig. 1). Compared with the control group (CN), ALFF’s T-value for both the T group and T+H group in the left and right dorsolateral Superior-Frontal Gyrus (SFG) were significantly lower than the global mean values from the control (CN) group (p < 0.05) (Table. 2a). A two-sample t-test did not reveal any statistical differences between the T group and T+H group in the left and right Dorsolateral Superior Frontal Gyrus (SFG) for ALFF analysis (p > 0.05).

**Table 2a.**
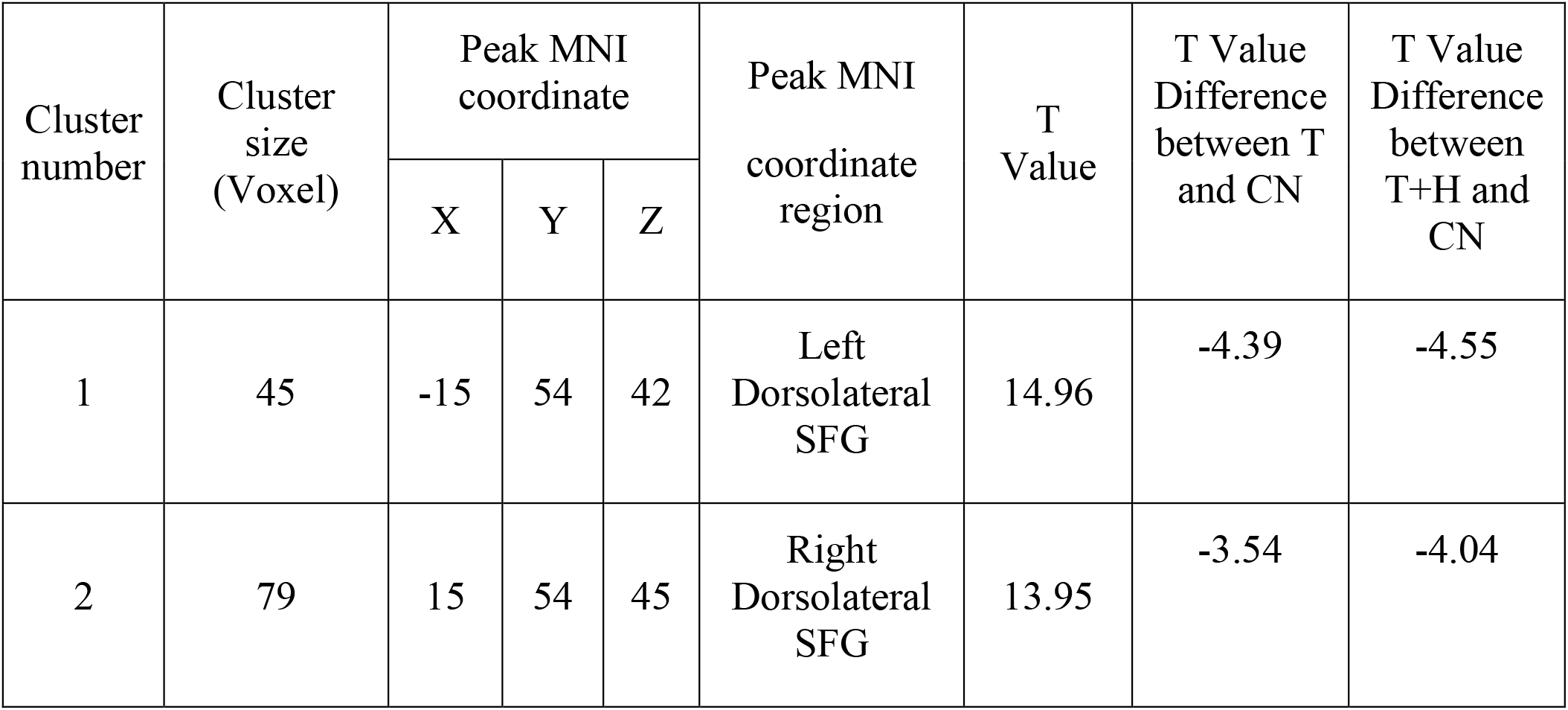
Decreased ALFF activities in both Tinnitus (T) and Tinnitus with Hearing loss (T+H) groups than in the control group (CN)

### 3.2 ReHo Results

We also discovered significant ReHo value differences in the right dorsolateral SFG for both T and T+H groups compared to the control (CN) group (Fig. 2). Regarding ReHo’s T-value, both the T group and T+H group in the right dorsolateral SFG revealed significantly lower values than the global mean values from the control (CN) group (p < 0.05) (Table. 2b). A two-sample t-test did not reveal any statistical differences between the T group and T+H group in the right Dorsolateral Superior Frontal Gyrus (SFG) (p > 0.05).

**Table 2b.**
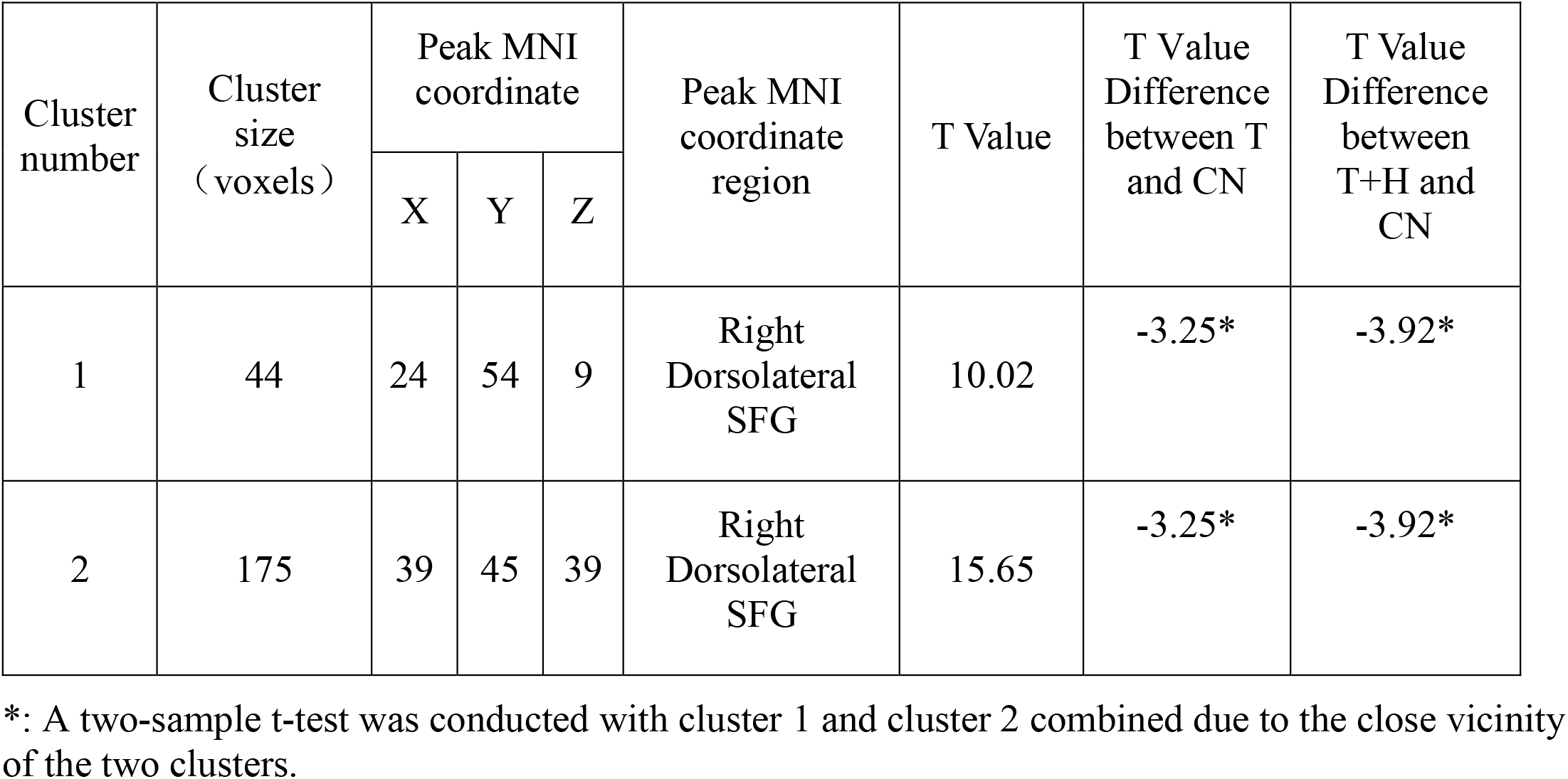
Decreased ReHo activities in both Tinnitus (T) and Tinnitus with Hearing loss (T+H) groups than in the control group (CN).

### 3.3 Voxel-Wise Functional Connectivity (FC) Results

Two regions identified from the ALFF analysis (dorsolateral SFG, left and right) were used as seeds for further FC analysis. Brain regions with significant functional connectivity pattern differences for the ALFF analysis clusters 1 and 2 were demonstrated in Figures 3 and 4, respectively. In contrast to the control group (CN), both the T group and T+H group exhibited a reduction in connectivity between the seed region in the left dorsolateral SFG (ALFF cluster 1) and right dorsolateral SFG, left medial Superior Frontal Gyrus (SFG), and right Superior Parietal Gyrus (SPG) (p < 0.05) (Table 3a). No difference was observed between the T and T+H groups (p > 0.05). At the same time, both the T group and T+H group exhibited decreased connectivity between the seed region in the right dorsolateral SFG (ALFF cluster 2) and right Middle Frontal Gyrus (MFG), right medial Superior Frontal Gyrus (SFG), and right Superior Parietal Gyrus (SPG) (p < 0.05) (Table 3b). No difference was observed between the T and T+H groups (p > 0.05).

**Table 3a.**
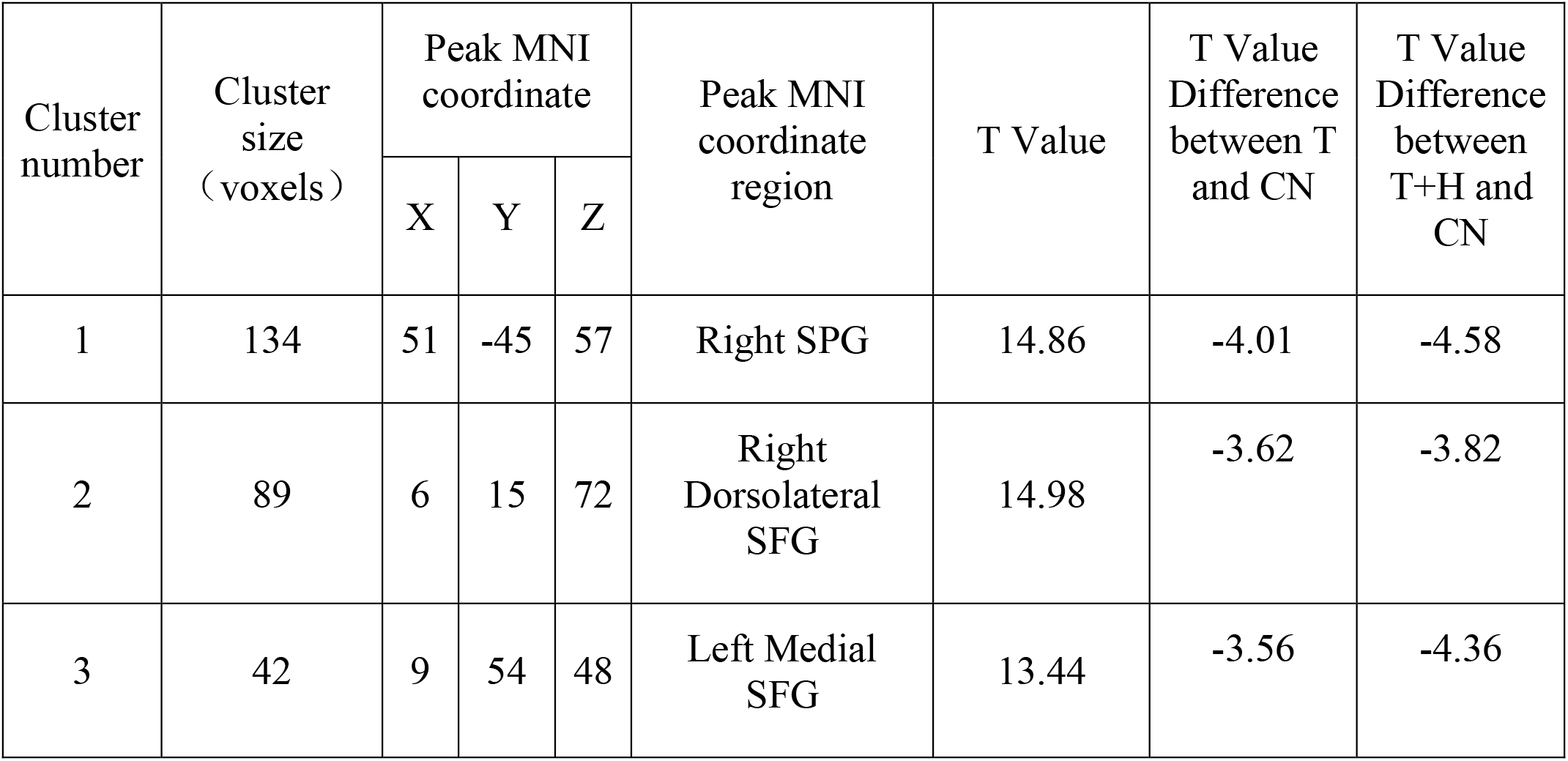
Decreased activities in Voxel-Wise Functional Connectivity (FC) ALFF cluster 1 for both Tinnitus (T) and Tinnitus with Hearing loss (T+H) groups than in the control group (CN)

**Table 3b.**
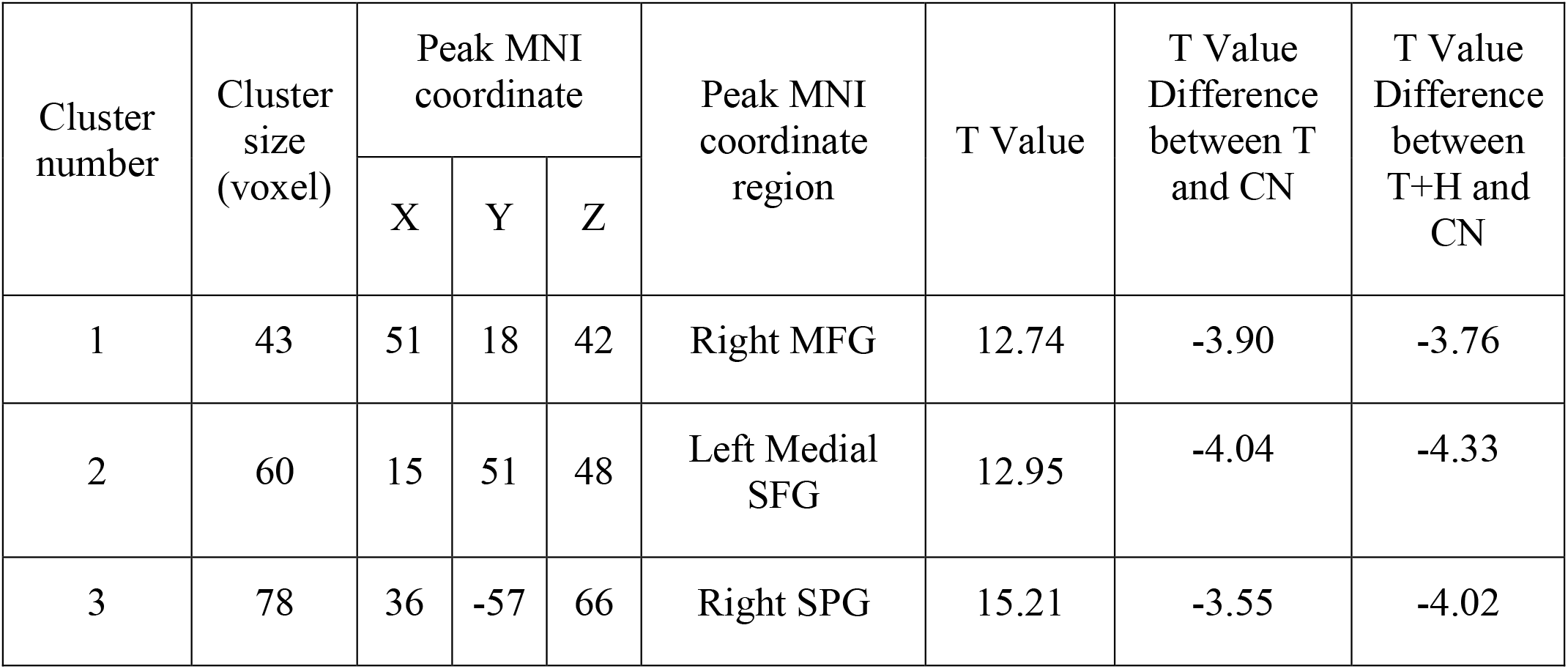
Decreased activities in Voxel-Wise Functional Connectivity (FC) ALFF cluster 2 for both Tinnitus (T) and Tinnitus with Hearing loss (T+H) groups than in the control group (CN)

Due to the close vicinity of clusters 1 and 2 identified from the ReHo analysis (dorsolateral SFG, right), we combined both clusters and used them as the seed region for further functional connectivity analysis. Brain regions with significant functional connectivity pattern differences were illustrated in Figure 5. In contrast to the control group (CN), both the T group and T+H group demonstrated lower connectivity levels between the seed region in the right dorsolateral SFG and left Superior Medial Frontal Gyrus (mSFG), right Middle Frontal Gyrus (MFG), and right Superior Parietal Gyrus (SPG) (p < 0.05) (Table 3c). However, we did not discover any significant difference between the T and T+H groups (p>0.05).

**Table 3c.**
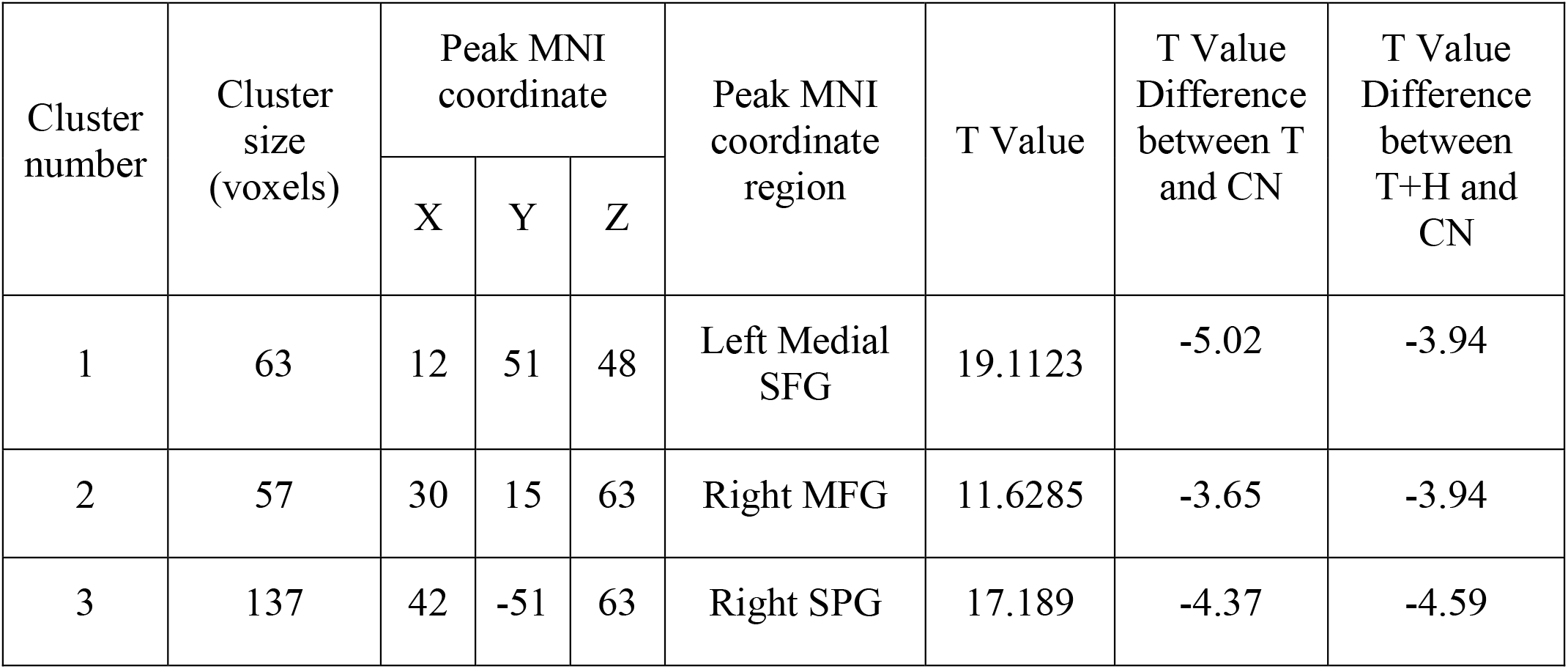
Decreased activities in Voxel-Wise Functional Connectivity (ReHo Cluster 1 and 2 combined) for both T and T+H groups than in the control group (CN)

## 4 Discussion

One major obstacle in uncovering tinnitus pathophysiology is that every tinnitus patient has a unique tinnitus profile. For example, while patients with hearing loss do not necessarily develop tinnitus, some patients with clinically normal hearing can perceive tinnitus. In addition, several brain networks associated with memory, attention, and emotional processing are involved in the tinnitus generation. Different levels of involvement among different brain regions and networks reflect the variable aspects of a patient’s tinnitus profile.

In the current study, we utilized various resting-state fMRI technologies, including the ALFF, ReHo, and Voxel-Wise functional connectivity (FC), to investigate the differences in the intra-regional brain activity and inter-regional functional connectivity in patients with tinnitus only (T) and tinnitus with hearing loss (T+H). Compared with the control group (CN), subjects from the T and T+H groups demonstrated decreased activity in the dorsolateral SFG (left and right) through the initial ALFF and ReHo analyses. In addition, the follow-up voxel-wise functional connectivity (FC) revealed decreased connection activity between the dorsolateral SFG (left and right) and right Superior Parietal Gyrus (SPG), right Middle Frontal Gyrus (MFG), and left medial Superior Frontal Gyrus (mSFG). These findings are in agreement with existing literature, which revealed that patients with hearing loss and tinnitus demonstrated abnormal intra-regional neural activity and disrupted connectivity in the hub regions of some non-auditory networks, including the default mode network (DMN), optical network, dorsal attention network (DAN), and central executive network (CEN). Nevertheless, we did not discover significant differences between the T and the T+H groups within these areas. This finding challenges the notion that neural plasticity associated with tinnitus differs from plasticity associated with tinnitus with hearing loss. As a result, we conclude that hearing loss might not be the tinnitus generator throughout our findings.

Furthermore, our findings suggested heavy involvement of brain regions responsible for attention and stimuli monitoring and orientations. Understanding their unique roles and involvement in attention and stimuli monitoring and orientations can help us better understand the mechanisms behind tinnitus generation.

### 4.1 Role of Dorsolateral Superior Frontal Gyrus (SFG) in Tinnitus Generation

The superior frontal gyrus (SFG) is critical for motor movement, working memory, resting state, and cognitive control. Human neuroimaging studies, anatomic experiments, and lesions have indicated that neoplastic changes or neuropsychiatric pathologies can disrupt the SFG’s functional processes (Boisgueheneuc et al., 2006; Kinoshita et al., 2012; Li et al., 2013). The dorsolateral SFG is responsible for top-down processing and cognitive functions, including working memory, episodic memory, goal-driven attention, planning, problem-solving, and task-switching. These findings support the dorsolateral SFG engagement in the CEN manipulations (Kinoshita et al., 2012; Hu et al., 2016).

In addition to the resting-state functional connectivity (rs-FC) with the middle frontal gyrus (MFG), the dorsolateral SFG was functionally correlated with the Default Mode Network (DMN), especially the precuneus. Existing literature describes the DMN as a classical ‘intrinsic’ system, specializing in internally-oriented cognitive processes such as conceptual processing, daydreaming, and future planning (Cloutman & Lambon Ralph, 2012; Lin et al., 2017).

Although the DMN and the CEN have been recognized as two functionally anti-correlated networks, studies revealed that they are responsible for dynamic interaction and efficient allocation of attention. The neural substrate for the functional interaction between the two networks has been observed from the posterior cingulate cortex (PCC) and the precuneus (Schmidt et al., 2013; Chen et al., 2018). Furthermore, existing studies discovered that memory tasks could simultaneously activate dorsolateral SFG and the PCC, suggesting their involvement in the CEN (Nagahama et al., 1999; Kinoshita et al., 2012; Schmidt et al., 2017). Therefore, we suggest that the dorsolateral SFG may serve as a regulation center between the CEN and DMN. When the dorsolateral SFG demonstrated reduced activity, the CEN could be in a disrupted state, leading to a reduced top-down attention-filtering capability. This theory can explain tinnitus patients’ typical report of difficulty filtering out tinnitus during quiet situations.

### 4.2 Role of Medial Superior Frontal Gyrus (mSFG) in Tinnitus Generation

Existing literature revealed that the medial SFG has anatomic connections with the cingulate cortex (mainly the anterior and medial section of the cingulate cortex, ACC & MCC) through the cingulum and that functional correlation with the MCC and the DMN (Nagahama et al., 1999). In addition, dense connections between the dorsolateral prefrontal cortex (DLPFC) (including the SFG) and the ACC and MCC have also been discovered in humans (Zhang et al., 2011; Cloutman & Lambon Ralph, 2012; Ueyama et al., 2013).

Moreover, the rs-FCs between the SFG, ACC, and MCC have been reported (Cloutman & Lambon Ralph, 2012; Yang et al., 2014; Khan et al., 2021). The anatomical and functional connections between the medial SFG and the anterior MCC suggest that the medial SFG is involved in cognitive control because the anterior part of the MCC has been related to cognitive control, including conflict monitoring, response selection, error detection, and attention manipulation. Additionally, the medial SFG demonstrates anatomic connections with the ACC, a core node of the DMN, and functional correlation with the DMN, suggesting that the medial SFG is a critical manipulation center for the DMN (Hu et al., 2021).

Therefore, we propose that reduced functional connectivity between the dorsolateral SFG and the medial SFG causes DMN regulation disruption, reducing patients’ ability to manipulate attention. This significant change within the top-down attention regulating mechanism leads to an increase in tinnitus perception, regardless of hearing loss.

### 4.3 Role of Right Middle Frontal Gyrus (MFG) in Tinnitus Generation

As a critical component of the ventral attention network (VAN), the right middle frontal gyrus (MFG) served as a convergence center for the DAN and the VAN by working as a circuit-breaker to interrupt ongoing endogenous attentional processes in the DAN and reorient attention to an exogenous stimulus (Japee et al., 2015; Briggs et al., 2021). Furthermore, the right MFG is active when reorienting to distinctive signals from unexpected locations (Carter et al., 2006).

Reduced functional connectivity between the dorsolateral SFG and the right MFG could disrupt the “circuit-breaker” between the VAN and the DAN, where the right MFG is no longer adequate for attention orientation to novel stimuli. From a clinical standpoint, this agrees with the typical description from the tinnitus patients that they unconsciously perceive their tinnitus to be more prominent in quieter situations, regardless of hearing loss.

### 4.4 Role of Superior Parietal Gyrus (SPG) in Tinnitus Generation

The SPG performs complex functions, including spontaneous attention regulation and top-down processing. Existing literature also revealed that the SPG became more active during a task-free resting state. As a critical component of the superior parietal lobule (SPL), the SPG links closely with the occipital lobe and involves the somatosensory and visuospatial integration, attention, written language, and working memory (Berlucchi & Vallar, 2018). Existing literature also reported SPG’s implications in shifting attention between visual targets and spatial-related attention shifts state (Lin et al., 2021). Thus, reduced functional connectivity between the dorsolateral SFG and the SPG could disrupt tinnitus patients’ working memory. This finding agrees with the typical complaint from the tinnitus patients that they have more difficulty filtering out the tinnitus without the presence of external stimuli.

### 4.5 Clinical Significance of Our Findings in Tinnitus Management

There is no resolute treatment for tinnitus, considering the cause can be multifactorial. However, clinicians can effectively manage tinnitus with the help of multidisciplinary options. According to the clinical practice guideline for tinnitus from the American Academy of Otolaryngology-Head and Neck Surgery, the primary tinnitus management options should include patient education and counseling, hearing amplification, sound therapy, and cognitive-behavioral therapy (CBT). The guideline did not recommend medical therapy, dietary supplements, and repetitive transcranial magnetic stimulation for tinnitus management (Tunkel et al., 2014; Zenner et al., 2016).

Our finding suggested that reduced activity levels within the brain regions responsible for top-down attention and stimuli monitoring and orientations contribute to tinnitus generation, regardless of hearing loss. This conclusion agrees with the existing literature that the impact of tinnitus on quality of life correlates more with psychological factors than acoustic properties of tinnitus. Therefore, from the clinical perspective, we can minimize the impact of tinnitus by re-calibrating the activities of these brain regions. The CBT can effectively achieve this goal by addressing the irrational and biased thought processes and behavioral and emotional dysfunctions that reduce patients’ quality of life (Fuller et al., 2020). In addition, clinicians can implement sound therapy to de-sensitize patients’ attention to tinnitus, especially in quiet environments (Liu et al., 2021; Osuji, 2021). Finally, clinicians can customize these management strategies for tinnitus patients, depending on patients’ symptoms.

## 5 Conclusion

Our project indicated a reduced activity level within the dorsolateral SFG (left and right) region, using ALFF and ReHo analyses. In addition, our follow-up Voxel-Wise functional connectivity revealed decreased connection activity between the dorsolateral SFG (left and right) and right Superior Parietal Gyrus (SPG), right Middle Frontal Gyrus (MFG), and left medial Superior Frontal Gyrus (mSFG). We did not find significant differences between tinnitus-only subjects and subjects with hearing loss and tinnitus. Our data suggested that disruptions in brain regions responsible for attention and stimuli monitoring and orientations contribute to tinnitus generation. Thus, hearing loss might not be the primary cause of tinnitus. We strongly recommend cognitive-behavioral therapy (CBT) and sound therapy for tinnitus management based on the evidence presented.

## Data Availability

All data produced in the present work are contained in the manuscript.

## Author Contributions

**Haoliang Du**^**1**^: Conceptualization, Methodology, Investigation, Formal analysis, Writing-Original Draft Preparation. **Xu Feng** ^**1†**^: Resources, Investigation, Data Curation, Writing - Review & Editing. **Xiaoyun Qian**^**2**^: Supervision, Project administration, Writing - Review & Editing, Funding acquisition. **Jian Zhang**^**3**^: Resources, Software, Validation, Visualization, Formal Analysis. **Bing Liu**^**4**^: Resources, Software, Validation, Formal Analysis. **Xia Gao*:** Supervision, Project administration, Funding acquisition. **Zhichun Huang*:** Supervision, Project administration, Funding acquisition

All authors contributed to the article and approved the submitted version.

## Acknowledgments

We would like to express our most sincere appreciation to Dr. Xia Gao and Dr. Zhichun Huang for their supervision, guidance, and encouragement throughout this project. We would also like to extend our gratitude to the Department of Otolaryngology-Head and Neck Surgery and the Department of Radiology at Nanjing Zhongda Hospital, affiliated with Southeast University, for graciously sharing precious clinical data with us. All authors are supported by The Project of Invigorating Health Care through Science, Technology, and Education.

