## Supplementary material for "Hearing Loss and Tinnitus: Uncover the Mechanism of Tinnitus using Resting-State Functional Magnetic Resonance Imaging (rs-fMRI) Technologies": Table

Tables

**Table 1**

Subject Characteristics of the Tinnitus Group (T), Tinnitus with Hearing Loss Group (T+H), and Control Group (CN)

|  | T Group | T+H Group | CN Group | p-value |
| --- | --- | --- | --- | --- |
| Age (Year) | 36.43±2.48 | 48.84±2.20 | 37.38±1.84 | >0.05 |
| Gender (Male: Female) | 12:9 | 16:16 | 20:9 | >0.05 |
| Education Duration (Year) | 9.56±2.42 | 9.88±1.86 | 10.12±2.43 | >0.05 |
| Tinnitus Duration (Year) | 3.44±1.08 | 2.25±0.99 | -- | >0.05 |
| Sleep Quality | 1.90±0.27 | 1.75±0.24 | -- | >0.05 |
| THQ Total Score | 49.27±4.28 | 40.01±3.96 | -- | >0.05 |
| SAS Score | 39.33±1.67 | 34.16±0.97 | -- | >0.05 |
| SDS Score | 39.48±2.36 | 37.09±1.67 | -- | >0.05 |

Data are represented as Mean ± SD

**Table 2a** Decreased ALFF activities in both Tinnitus (T) and Tinnitus with Hearing loss (T+H) groups than in the control group (CN)

| Cluster number | Cluster size (Voxel) | Peak MNI coordinate | | | Peak MNI  coordinate region | T Value | T Value Difference between T and CN | T Value Difference between T+H and CN |
| --- | --- | --- | --- | --- | --- | --- | --- | --- |
|  |  | X | Y | Z |  |  |  |  |
| 1 | 45 | -15 | 54 | 42 | Left Dorsolateral SFG | 14.96 | -4.39 | -4.55 |
| 2 | 79 | 15 | 54 | 45 | Right Dorsolateral SFG | 13.95 | -3.54 | -4.04 |

**Table 2b** Decreased ReHo activities in both Tinnitus (T) and Tinnitus with Hearing loss (T+H) groups than in the control group (CN).

| Cluster number | Cluster size（voxels） | Peak MNI coordinate | | | Peak MNI coordinate region | T Value | T Value Difference between T and CN | T Value Difference between T+H and CN |
| --- | --- | --- | --- | --- | --- | --- | --- | --- |
|  |  | X | Y | Z |  |  |  |  |
| 1 | 44 | 24 | 54 | 9 | Right Dorsolateral SFG | 10.02 | -3.25* | -3.92* |
| 2 | 175 | 39 | 45 | 39 | Right Dorsolateral SFG | 15.65 | -3.25* | -3.92* |

*: A two-sample t-test was conducted with cluster 1 and cluster 2 combined due to the close vicinity of the two clusters.

**Table 3a** Decreased activities in Voxel-Wise Functional Connectivity (FC) ALFF cluster 1 for both Tinnitus (T) and Tinnitus with Hearing loss (T+H) groups than in the control group (CN)

| Cluster number | Cluster size（voxels） | Peak MNI coordinate | | | Peak MNI coordinate region | T Value | T Value Difference between T and CN | T Value Difference between T+H and CN |
| --- | --- | --- | --- | --- | --- | --- | --- | --- |
|  |  | X | Y | Z |  |  |  |  |
| 1 | 134 | 51 | -45 | 57 | Right SPG | 14.86 | -4.01 | -4.58 |
| 2 | 89 | 6 | 15 | 72 | Right Dorsolateral SFG | 14.98 | -3.62 | -3.82 |
| 3 | 42 | 9 | 54 | 48 | Left Medial SFG | 13.44 | -3.56 | -4.36 |

**Table 3b** Decreased activities in Voxel-Wise Functional Connectivity (FC) ALFF cluster 2 for both Tinnitus (T) and Tinnitus with Hearing loss (T+H) groups than in the control group (CN)

| Cluster number | Cluster size (voxel) | Peak MNI coordinate | | | Peak MNI coordinate region | T Value | T Value Difference between T and CN | T Value Difference between T+H and CN |
| --- | --- | --- | --- | --- | --- | --- | --- | --- |
|  |  | X | Y | Z |  |  |  |  |
| 1 | 43 | 51 | 18 | 42 | Right MFG | 12.74 | -3.90 | -3.76 |
| 2 | 60 | 15 | 51 | 48 | Left Medial SFG | 12.95 | -4.04 | -4.33 |
| 3 | 78 | 36 | -57 | 66 | Right SPG | 15.21 | -3.55 | -4.02 |

**Table 3c** Decreased activities in Voxel-Wise Functional Connectivity (ReHo Cluster 1 and 2 combined) for both T and T+H groups than in the control group (CN)

| Cluster number | Cluster size (voxels) | Peak MNI coordinate | | | Peak MNI coordinate region | T Value | T Value Difference between T and CN | T Value Difference between T+H and CN |
| --- | --- | --- | --- | --- | --- | --- | --- | --- |
|  |  | X | Y | Z |  |  |  |  |
| 1 | 63 | 12 | 51 | 48 | Left Medial SFG | 19.1123 | -5.02 | -3.94 |
| 2 | 57 | 30 | 15 | 63 | Right MFG | 11.6285 | -3.65 | -3.94 |
| 3 | 137 | 42 | -51 | 63 | Right SPG | 17.189 | -4.37 | -4.59 |
