## Supplementary material for "Hearing Loss and Tinnitus: Uncover the Mechanism of Tinnitus using Resting-State Functional Magnetic Resonance Imaging (rs-fMRI) Technologies": Figure

Figures

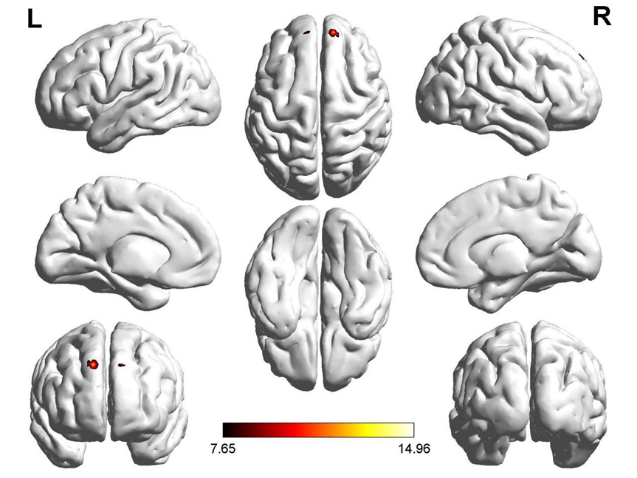
**Fig. 1.** Significant ALFF value differences in the left and right dorsolateral SFG for both Tinnitus (T) and Tinnitus with Hearing loss (T+H) groups (P <0.001 corrected for multiple comparisons using FamilyWise Error Correction).

**Fig. 2.** Significant ReHo value differences in the right dorsolateral SFG for both Tinnitus (T) and Tinnitus with Hearing loss (T+H) groups, compared to the CN group (P <0.001 corrected for multiple comparisons using FamilyWise Error Correction)

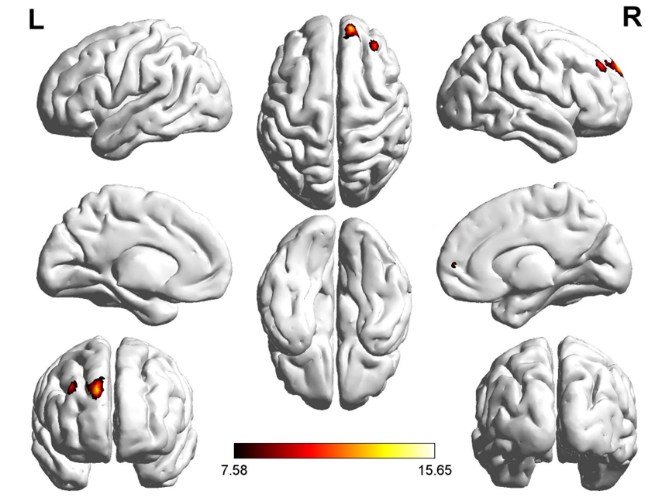

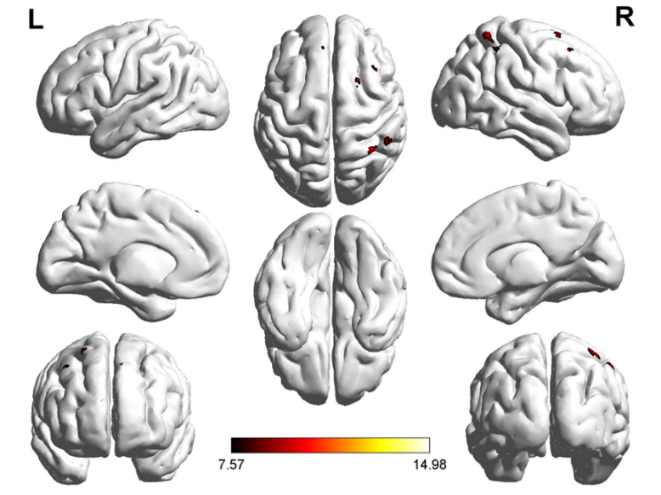
**Fig. 3** Significant FC patterns between the ROIs (dorsolateral SFG, left and right) and the right SFG, left medial Superior Frontal Gyrus (SFG), and right Superior Parietal Gyrus (SPG). Significant thresholds were corrected by using Family-Wise Error (FEW) criterion and set at p<0.001.

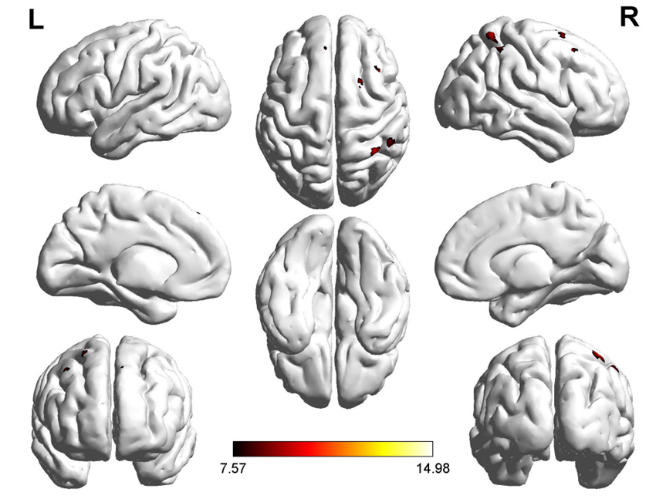
**Fig. 4.** Significant FC patterns between the left and right dorsolateral SFG and the right Middle Frontal Gyrus (MFG), right medial Superior Frontal Gyrus (SFG), and right Superior Parietal Gyrus (SPG) in the whole brain by using one-sample t-test in both T group and T+H group. Significant thresholds were corrected by using Family-Wise Error (FEW) criterion and set at p<0.001.

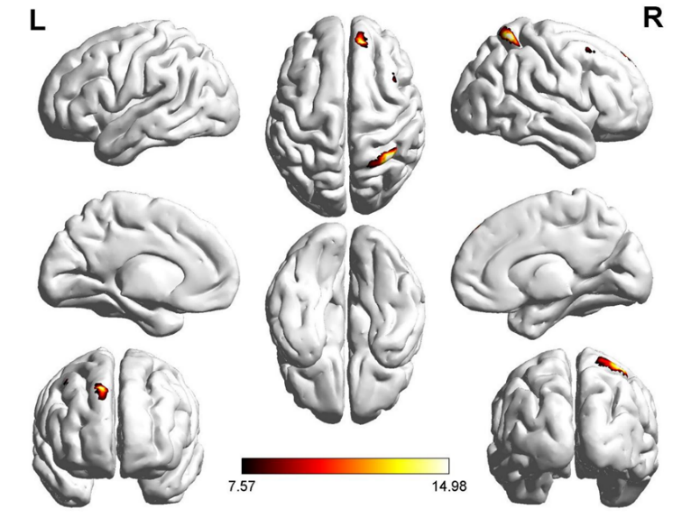
**Fig. 5.** Significant FC patterns between the right dorsolateral SFG and the left Medial Superior Frontal Gyrus (mSFG), Right Middle Frontal Gyrus (MFG), and right Superior Parietal Gyrus (SPG). Significant thresholds were corrected by using the FDR criterion and set at p<0.001.
